# Prenatal cannabis exposure is associated with localized brain differences that partially mediate associations with increased adolescent psychopathology

**DOI:** 10.1101/2023.09.19.23295792

**Authors:** David AA Baranger, Alex P Miller, Aaron J Gorelik, Sarah E Paul, Alexander S Hatoum, Emma C Johnson, Sarah MC Colbert, Christopher D Smyser, Cynthia E Rogers, Janine D Bijsterbosch, Arpana Agrawal, Ryan Bogdan

## Abstract

Prenatal cannabis exposure (PCE) is associated with mental health problems, but the neurobiological mechanisms remain unknown. We find that PCE is associated with localized differences across neuroimaging metrics that longitudinally mediate associations with mental health in adolescence (n=9,322-10,186). Differences in brain development may contribute to PCE-related variability in adolescent mental health.

## Main

Alongside increasingly permissive sociocultural attitudes and laws, cannabis use during pregnancy doubled between 2002 (3.4%) and 2017 (7%)^1^, despite evidence of potential adverse consequences and discouragement from governmental health agencies (e.g., Surgeon General, Food and Drug Administration)^2,3^ and professional organizations (American College of Obstetricians and Gynecologists)^4^. Accumulating studies link prenatal cannabis exposure (PCE) to adverse behavioral outcomes during childhood, adolescence, and early adulthood (e.g., increased psychopathology, reduced cognition^5–8^; see also^9–11^), suggesting that PCE may influence brain development. As cannabis constituents traverse the placenta^12^ and interface with the fetal endocannabinoid system, which critically contributes to neurodevelopment (e.g., axonal elongation, synaptic plasticity, synaptic pruning)^13^, there are plausible molecular mechanisms through which PCE may impact brain development. However, there has been a dearth of research examining such putative neural system level mechanisms^14^, which are needed to appropriately evaluate the safety of cannabis use during pregnancy.

Using data from the Adolescent Brain Cognitive Development^SM^ (ABCD®) Study^15^ (data release 5.0) of 11,875 children, we tested whether PCE before and after maternal knowledge of pregnancy is associated with multimodal brain metrics. These included resting-state fMRI (rs-fMRI) cortical and subcortical connectivity, volume, surface area, thickness, and sulcal depth, and measures from diffusion tensor imaging (DTI) and restriction spectrum imaging (RSI) models applied to diffusion weighted data from white matter tracts, cortical white matter, and gray matter. Notably, endocannabinoid receptors are not expressed in the fetus until 5 to 6 weeks’ gestation^16–18^, which approximately corresponds to when, in this study, mothers learned they were pregnant (mean [SD], 6.9 [6.8] weeks). Thus, we hypothesized that the strength of associations with cannabis exposure would be stronger among children with PCE after maternal knowledge of pregnancy. Further, we test whether associated brain metrics were, in turn, associated with childhood psychopathology, and whether associations with psychopathology may be partially mediated by brain differences.

Analyses included data from the first two waves of neuroimaging data collection (baseline visit and follow-up wave 2), at ages 9-10 and 11-12. This comprised 16,641 observations from 10,186 participants who had complete usable data for at least one neuroimaging modality (6,455 participants had two data points available, 3,731 had one), including 373 prenatally exposed to cannabis only prior to the mother’s knowledge of her pregnancy (pre-knowledge only; ns = 337-373 per modality) and 195 exposed both before and after knowledge of her pregnancy (pre- and post-knowledge; ns = 172-195 per modality) (**Table S1, Online Methods; Supplemental Data**).

Linear mixed-effect models (**Online Methods**) revealed three brain metrics significantly associated with PCE after multiple-test correction for all tests (n_brain variables_=2,907; **Figure 1**; **Supplemental Data**): 1) restricted normalized directional diffusion of the forceps minor, 2) transverse diffusivity of the right pars triangularis, and 3) rs-fMRI connectivity of the auditory network and left putamen. An additional 14 brain metrics were significantly associated with PCE after multiple-test correction within modality (**Figure 1**; **Figure 2**; **Supplemental Data**). These broadly included diffusion weighted metrics from gray and white matter, and white matter tracts, in the frontal and parietal lobes. Findings indicate that the association of PCE with brain metrics is relatively localized; no global measure (**Online Methods**) survived correction (smallest p=0.015 uncorrected). While several associations were, as hypothesized, driven by stronger effects in the pre- and post-knowledge group (**Figure 2**; e.g., transverse and mean diffusivity of the cortical gray matter in the right pars triangularis), there were also associations driven by stronger effects in the pre-knowledge group (**Figure 2**; e.g., hindered and restricted normalized directional diffusion in the cortical white matter of the inferior frontal cortex). Notably, regions where effects were driven by the pre-knowledge group did not show associations with psychopathology in subsequent analyses. All findings were robust to additional *post-hoc* tests, including: 1) inclusion of pregnancy-related variables with high missingness (due to non-report); 2) restricting data to only the baseline visit; 3) adjustment for polygenic risk for cannabis use disorder (**Online Methods; Supplemental Data)**.

**Figure 1.**
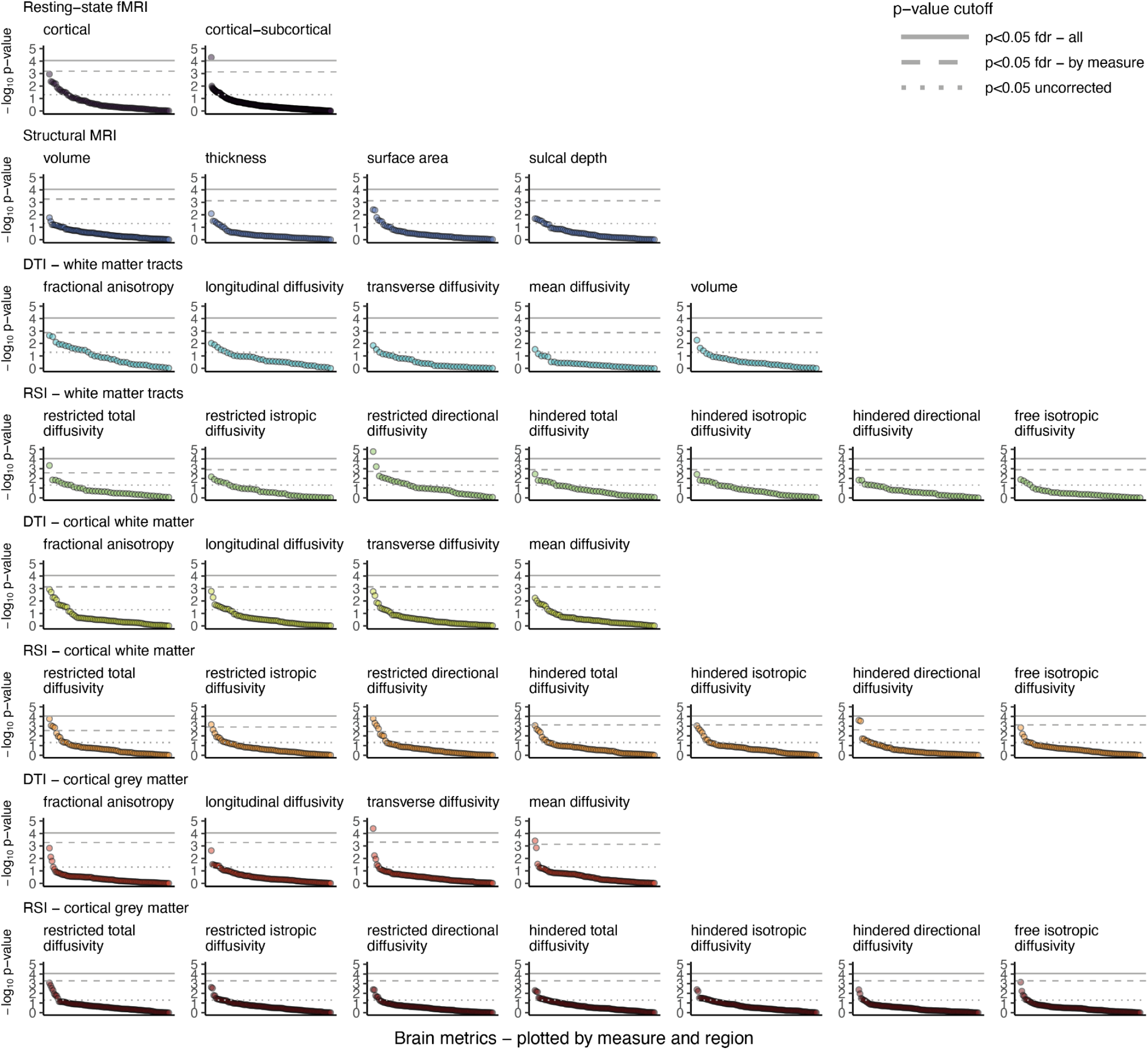
Association of prenatal cannabis exposure with brain metrics. Plots show the -log10 (negative base-10 logarithm) of the p-value for the association between brain metrics and prenatal cannabis exposure (PCE) from mixed effect regressions. Each imaging modality is plotted in a separate plot, with regions ordered within each plot from most to least significant. Horizontal lines are placed at p=0.05 uncorrected (dotted gray line), p<0.05 fdr-corrected for all comparisons of a given measure (i.e., all metrics within one plot; dashed gray line), and p<0.05 fdr-corrected for all 2,907 tests (solid gray line). **Figure 2** lists all regions surviving fdr-correction for comparisons of a given measure. Additional statistical information is provided in the **Supplemental Data File**.

**Figure 2.**
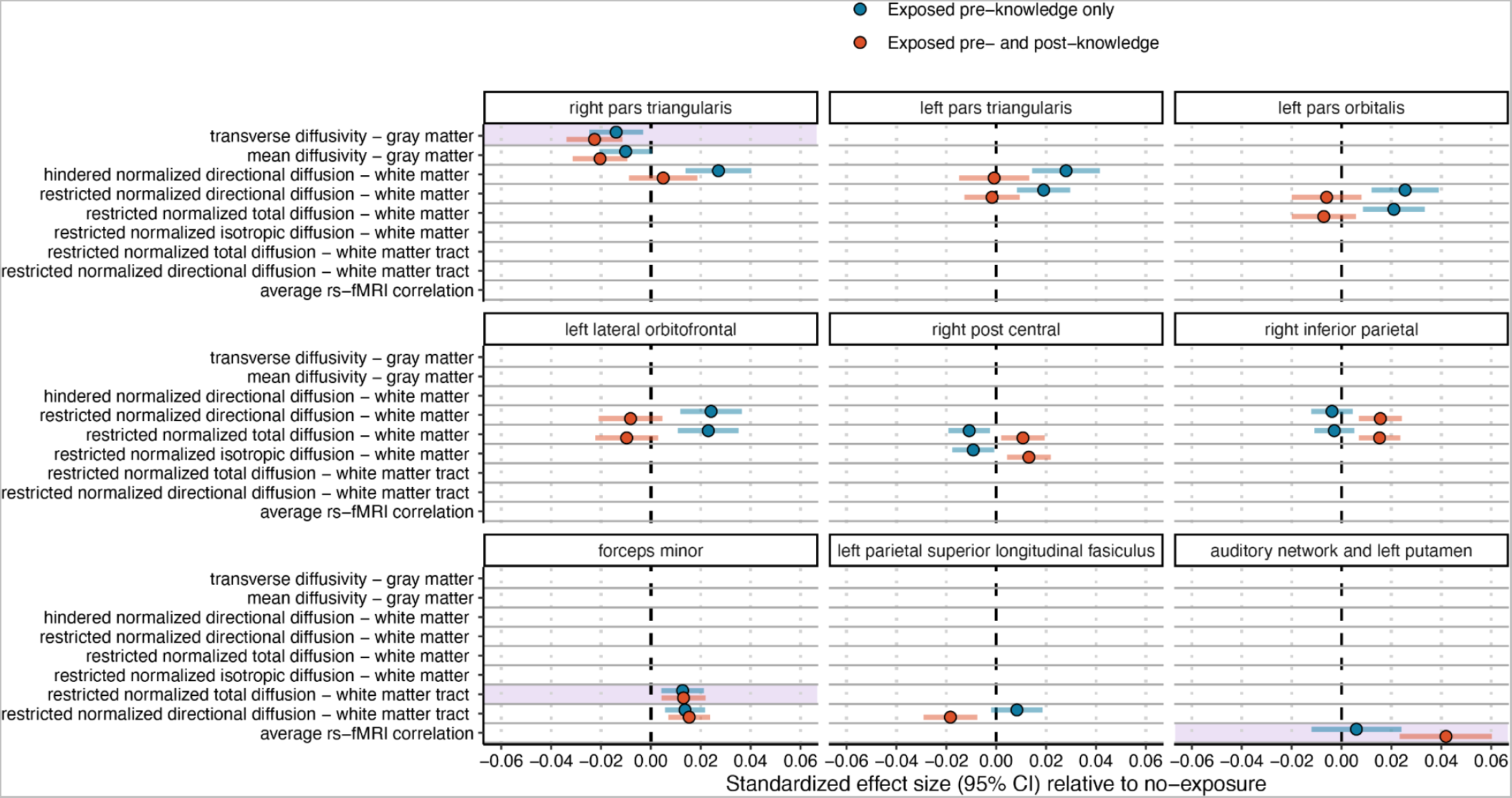
Significant associations of prenatal cannabis exposure with brain metrics, by exposure group. Standardized regression β effect sizes and 95% CIs from mixed-effect regressions assessing the association of prenatal cannabis exposure prior to maternal knowledge of pregnancy or prior to and post-maternal knowledge of pregnancy compared with no exposure. Nonsignificant outcomes are not shown. Y-axis reflects the measure-type, and each plot shows a single brain region to highlight overlap across different measures. Note that not all regions are measured with all modalities. Additional statistical information is provided in the **Supplemental Data File**. Regions surviving FDR multiple-test correction for all 2,907 tests are highlighted in purple.

To evaluate whether the 17 significant PCE-brain associations may plausibly contribute to behavioral variability, we estimated their associations with 13 measures of adolescent mental health that have previously been associated with PCE in this sample (**Figure S1**)^19–21^, including measures reflecting psychotic-like experiences, aggressive behavior, attention problems, and social problems (**Online Methods**). Seven associations survived multiple-test correction for all 221 tests, of which five remained significant when controlling for all PCE-related covariates used in primary analyses above; an additional eleven survived multiple-test correction within measure-type, of which six remained significant when controlling for additional PCE-related covariates (**Figure 3A**; **Supplemental Data; Online Methods**). Significant associations were all externalizing-related behaviors (i.e., attention and conduct problems, ADHD, and rule-breaking behavior) and were restricted to four metrics (**Figure 3A**) - including two which survived correction for all tests in the above analyses of associations with PCE - the right pars triangularis (transverse and mean diffusivity of the cortical gray matter) and the forceps minor (restricted directional and total diffusion). The directions of these associations are consistent with these brain regions mediating the effect of PCE on childhood psychopathology, including that the association of PCE with these metrics is driven by the pre- and post-knowledge group. Longitudinal mediation analysis (**Online Methods**) of these 11 brain-behavior associations revealed 8 indirect associations that were robust to multiple testing correction, of which 7 remained significant when controlling for additional PCE-related covariates (**Figure 3B; Supplemental Data; Online Methods**), wherein brain metrics partially mediated the association between PCE and attention and ADHD problems at a subsequent wave. Effects were uniformly small, indicating that 1.5-2% of the association of PCE with attention-related psychopathology may be plausibly mediated by these brain metrics.

**Figure 3.**
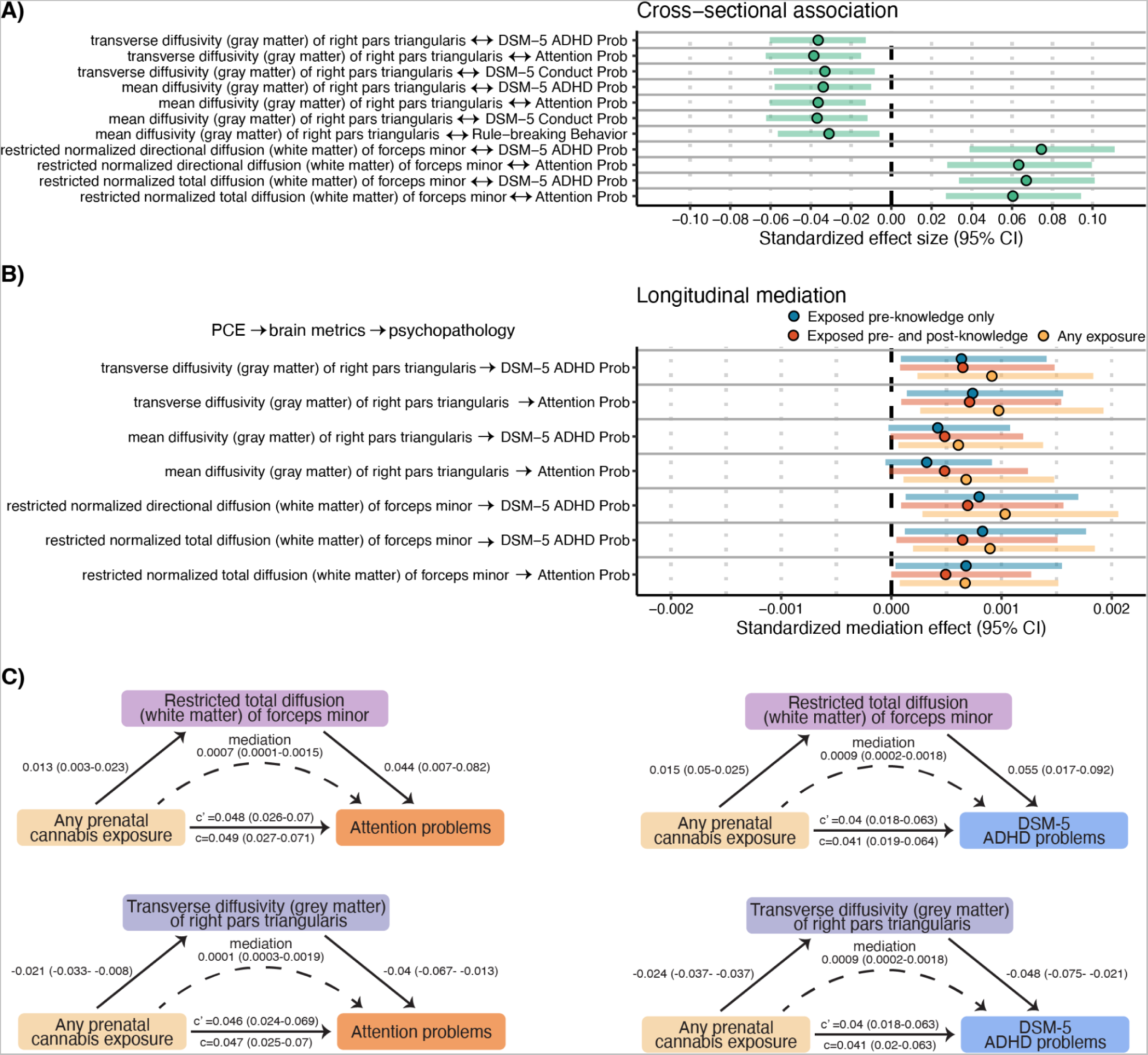
Association of brain metrics with childhood psychopathology. A) Standardized regression β effect sizes and 95% CIs from mixed-effect regressions assessing the association of significant (pfdr<0.05) brain metrics with psychopathology. B) Standardized mediation effect and 95% quasi-Bayesian CIs from longitudinal mediation analyses testing whether significant brain metrics mediate the association of prenatal cannabis exposure (PCE) with psychopathology. Analyses were run first collapsing across PCE groups (i.e., any exposure) and *post-hoc* analyses computed effects for each group separately. C) Mediation path diagrams for significant mediations (p<0.05 fdr) with regions which survived correction for all comparisons in primary analyses (**Figure 2**; restricted total diffusion of the forceps minor and transverse diffusivity of the right pars triangularis grey matter). Values reflect standardized effects and 95% CIs. c = total effect, c’ = direct effect.

These findings align with the hypothesis that the association of PCE with psychopathology may be partially attributable to effects of PCE on brain development. Our results build upon prior work using small sample sizes and a restricted subset of measures and regions^20,22–24^ to show that associations of PCE with brain metrics are relatively focal. In particular, we find robust associations with lower transverse and mean diffusivity of the right pars triangularis cortical gray matter and greater restricted directional and total diffusion of the forceps minor, both of which are, in turn, associated with attention-related psychopathology. An equivocal literature has inconsistently linked cannabis involvement in adults and adolescents (e.g., use, problematic use) to variability in other metrics within these regions (e.g., forceps minor fractional anisotropy, pars triangularis thickness)^25,26^. Our findings may reflect differential gray matter cellular organization (e.g., neurite density) and white matter tract integrity of these regions in participants with PCE. Physiological processes that increase isotropic diffusion (i.e., inflammation) would be expected to result in increased mean and transverse cortical diffusivity, and decreased restricted directional and total diffusion of white matter bundles^27,28^. Thus, our findings may reflect relatively reduced neuroinflammation in PCE participants that may begin in the intrauterine environment^29,30^, consistent with evidence that cannabinoids reduce the inflammatory response^31^. Alternatively, cortical mean diffusivity decreases over adolescence and restricted directional diffusion increases over adolescence^32^, and thus these results may indicate accelerated neurodevelopment with PCE, consistent with prior findings that chronic cannabis use is associated with accelerated aging^33^, possibly due to co-occurring exposure to post-combustion hydrocarbons^34^.

PCE has been broadly associated with psychopathology (e.g., 13 of 21 measures)^19^, yet the present results suggest that brain metrics account for a relatively modest portion of the association with a subset of externalizing-related measures. However, as hypothesized, the strongest effects were driven by participants with exposure both pre- and post-knowledge of pregnancy. This suggests that the outcome of PCE may depend on the developmental timing of exposure, and that improved measurement of the timing, mode, quantity, and frequency of exposure could yield larger effects.

While the ABCD study is among the largest studies of prenatal exposures and neurobiology, limitations include the relatively small sample of prenatal cannabis-exposed offspring. It should be noted that the measure of prenatal cannabis exposure used here reflects a retrospective report of behavior that occurred approximately 10 years earlier, which may have resulted in biased reporting and misclassification. Further, there is limited or no data on potency, mode, frequency, timing, or quantity of cannabis exposure in this data set. While we were able to account for many known familial, pregnancy-related, and child-related confounding variables, the role of unmeasured confounders (e.g., maternal stress during pregnancy) cannot be discounted. Mediation analyses show that a mediating effect of brain metrics is statistically plausible, but we cannot rule out reciprocal effects (i.e., psychopathology leads to changes in brain metrics) or unmeasured causal variables that drive changes in both brain metrics and psychopathology, among other explanations^35^.

Limitations notwithstanding, our study provides evidence that PCE is associated with differences in brain development that may partially mediate associations between PCE and increased adolescent psychopathology. Future work incorporating improved measurement of prenatal cannabis exposure, and exploring models which aggregate across multiple brain metrics, may yield larger effects and more insight into the causal mechanisms underlying these associations.

## Data availability

Data used in the preparation of this article were obtained from the Adolescent Brain Cognitive Development^SM^ (ABCD) Study (https://abcdstudy.org), held in the NIMH Data Archive (NDA). This is a multisite, longitudinal study designed to recruit more than 10,000 children aged 9-10 and follow them over 10 years into early adulthood. The ABCD Study® is supported by the National Institutes of Health and additional federal partners under award numbers U01DA041048, U01DA050989, U01DA051016, U01DA041022, U01DA051018, U01DA051037, U01DA050987, U01DA041174, U01DA041106, U01DA041117, U01DA041028, U01DA041134, U01DA050988, U01DA051039, U01DA041156, U01DA041025, U01DA041120, U01DA051038, U01DA041148, U01DA041093, U01DA041089, U24DA041123, U24DA041147. A full list of supporters is available at https://abcdstudy.org/federal-partners.html. A listing of participating sites and a complete listing of the study investigators can be found at https://abcdstudy.org/consortium_members/. ABCD consortium investigators designed and implemented the study and/or provided data but did not necessarily participate in the analysis or writing of this report. This manuscript reflects the views of the authors and may not reflect the opinions or views of the NIH or ABCD consortium investigators. The ABCD data repository grows and changes over time. The ABCD data used in this report came from http://dx.doi.org/10.15154/8873-zj65. DOIs can be found at https://nda.nih.gov/abcd/abcd-annual-releases.html. Dataset identifier: http://dx.doi.org/10.15154/dxx6-fk12.

## Acknowledgements

This study was supported by R01DA54750 (RB, AA). Additional funding included: DAAB (K99AA030808), APM (T32DA015035), AJG (DGE-213989), SEP (F31AA029934), ASH (K01AA030083), ECJ (K01DA051759; BBRF Young Investigator Grant 29571), CER (R01DA046224), AA (R01DA54750), RB (R01DA54750, R21AA027827, U01DA055367). Data for this study were provided by the Adolescent Brain Cognitive Development (ABCD) study which was funded by awards U01DA041022, U01DA041025, U01DA041028, U01DA041048, U01DA041089, U01DA041093, U01DA041106, U01DA041117, U01DA041120, U01DA041134, U01DA041148, U01DA041156, U01DA041174, U24DA041123, and U24DA041147 from the NIH and additional federal partners (https://abcdstudy.org/federal-partners.html). We thank Dr. Tayler Sheahan for her assistance with figure graphics.

## Online Methods

### MRI Acquisition and Processing

Casey et al., 2018^15^ provide an in-depth description of the ABCD Study^®^ imaging acquisition protocol and parameters and Hagler et al., 2019^36^ provide an in-depth description of the ABCD Study^®^ image processing and analysis methods^32^. The present analyses used tabulated neuroimaging data provided as part of the 5.0 data release. Here we provide a brief summary:

#### Structural MRI

1 mm isotropic T1-weighted structural magnetic resonance images (MRI) were acquired on 3 T (Siemens, Phillips and GE) MRI scanners using either a 32-channel head or 64-channel head-and-neck coil. Scan protocols were carefully harmonized across the three MRI vendor platforms to reduce scanner-caused variability. MRI data were processed with the Multi-Modal Processing Stream software package that includes FreeSurfer 5.3. Besides a modified intensity normalization process used by the ABCD processing pipeline, the standard FreeSurfer cortical and subcortical reconstruction pipeline was run to generate structural measures including volume, cortical thickness, cortical surface area, and cortical sulcal depth. Here we use measures from the Desikan cortical atlas and the Freesurfer Aseg subcortical atlas. A description of the quality-control measures conducted on the processed data is provided in Hagler et al. MRI analyses included only participants whose structural MRI reconstructions passed QC tests. Global variables used as covariates in analyses included intracranial volume, mean thickness, total surface area, and mean sulcal depth.

#### Diffusion MRI

Diffusion MRI (dMRI) data were acquired in the axial plane at 1.7 mm isotropic resolution with multiband acceleration factor 3. Diffusion-weighted images were collected with: seven b= 0 s/mm^2^ frames and 96 non-collinear gradient directions, 6 directions at b= 500 s/mm^2^, 15 directions at b= 1000 s/mm^2^, 15 directions at b= 2000 s/mm^2^, and 60 directions at b= 3000 s/mm^2^. 3D T2-weighted fast spin echo with variable flip angle scans were acquired at 1 mm isotropic resolution with no multiband acceleration. Scanning protocols were harmonized across sites.

Data were corrected for eddy current distortion. Images were rigid-body-registered to the corresponding volume synthesized from a robust tensor fit. Dark slices caused by abrupt head motion were replaced with values synthesized from the robust tensor fit, and the diffusion gradient matrix was adjusted for head rotation. Spatial and intensity distortions caused by B0 field inhomogeneity were corrected and gradient nonlinearity distortions were corrected for each frame. Data were registered to T1w structural images and dMRI data were then resampled to 1.7 mm isotropic resolution. Major white matter tracts were labeled using AtlasTrack. The diffusion tensor imaging (DTI) model was used to calculate standard measures related to microstructural tissue properties, including fractional anisotropy and mean, longitudinal (or axial), and transverse (or radial) diffusivity (MD, LD, and TD).

A Restriction Spectrum Imaging (RSI) model was also fit. This linear estimation approach allows for mixtures of “restricted” and “hindered” diffusion within individual voxels. RSI was used to model two volume fractions, representing intracellular (restricted) and extracellular (hindered) diffusion, with separate fiber orientation density (FOD) functions, modeled as fourth order spherical harmonic functions, allowing for multiple diffusion orientations within a single voxel. Measures derived from the RSI model fit include: restricted normalized isotropic, restricted normalized directional, restricted normalized total, hindered normalized isotropic, hindered normalized directional, hindered normalized total, and free normalized isotropic. Normalized isotropic and hindered normalized total reflect varying contributions of intracellular and extracellular spaces to isotropic diffusion-related signal decreases in a given voxel. Restricted normalized directional (a proxy for oriented myelin organization) and hindered normalized directional reflect oriented diffusion--diffusion that is greater in one orientation than others. Restricted normalized directional is similar to FA, except that it is unaffected by crossing fibers^32^. Restricted normalized total and hindered normalized total reflect the overall contribution to diffusion signals of intracellular and extracellular spaces.

Mean DTI and RSI measures were calculated for white matter fiber tract ROIs created with AtlasTrack and for ROIs derived from FreeSurfer’s automated subcortical segmentation. DTI and RSI measures were also sampled onto the FreeSurfer-derived cortical surface mesh to make maps of diffusion properties for cortical gray matter and white matter adjacent to the cortex. Here we use measures of these properties taken from the Desikan cortical atlas. dMRI analyses included only participants whose structural MRI and dMRI reconstructions passed QC tests. Global variables used as covariates in analyses included the mean of all regions/tracts for a given metric.

#### Resting-state fMRI

Twenty minutes of resting-state functional MRI (rs-fMRI) data were collected across four 5-minute scans with eyes open and passive viewing of a cross hair. The FIRMM real-time head motion monitoring system was implemented for motion detection in resting state fMRI scans at sites using Siemens scanners^37^. FIRMM allows scanner operators to adjust the scanning paradigm based on a participant’s degree of head motion (i.e., the worse the motion, the less usable data and greater the need for more data to be acquired). rs-fMRI data were acquired with: a matrix of 90 × 90, 60 slices, FOV size = 216 × 216, voxel size = 2.4 × 2.4 × 2.4 mm^3^, TR = 800 ms, TE = 30 ms, FA = 52°, and a multiband acceleration factor of 6. Head motion was further corrected by registering each frame to the first and images were corrected for distortions due to gradient nonlinearities. To correct for between-scan motion, each scan was resampled with cubic interpolation into alignment with a reference scan that is chosen as the one nearest to the middle of the set of fMRI scans for a given participant. Further processing included the removal of initial frames, normalization, regression of motion and mean signal time courses, and temporal filtering. Time points with a framewise displacement greater than 0.2 mm were excluded, as were periods with fewer than five contiguous, sub-threshold time points. Preprocessed time courses were sampled onto the cortical surface for each individual subject and average time courses were calculated for cortical surface-based ROIs using a functionally-defined parcellation based on resting-state functional connectivity patterns from the Gordon atlas. Average time courses were also calculated for subcortical ROIs. The correlation between each pair of ROIs were averaged within or between networks to provide summary measures of network correlation strength. rs-fMRI analyses included only participants whose data passed QC tests. Analyses of resting-state data did not include a global variable as a covariate.

### Measures of psychopathology

CBCL: The Child Behavior Checklist (CBCL)^38^ is a 113-item questionnaire on which caregivers rated items representing specific problems in the past six months. The CBCL was completed annually. Subscales include aggressive behavior, anxious/depressed, attention problems, rule-breaking behavior, somatic complaints, social problems, stress problems, thought problems, and withdrawn/depressed. There is a broadband scale for internalizing problems, which sums the anxious/depressed, withdrawn-depressed, and somatic complaints scores, and a broadband scale for externalizing problems, which sums rule-breaking and aggressive behavior. The total problems score is the sum of the scores of all the problem items. The CBCL also includes a set of DSM-oriented scales for depressive problems, affective problems, anxiety problems, somatic problems, ADHD problems, oppositional defiant problems, conduct problems, obsessive-compulsive problems, and sluggish cognitive tempo. Analyses using the CBCL included 12 scales which were previously found to be significantly associated with PCE in this sample (**Supplemental Figure 1**)^19^: Total problems, externalizing factor, rulebreaking behavior, aggressive behavior, social problems, thought problems, attention problems, sluggish cognitive tempo, stress problems, obsessive compulsive problems, ADHD problems, and conduct problems.

Psychotic-like Experiences: Children completed the 21-item Prodromal Questionnaire-Brief Child Version (PQ-BC)^39^ in which they are first asked to respond (yes/no) to whether they experienced a thought/feeling/experience (e.g., do familiar surroundings sometimes seem strange, confusing, threatening, or unreal to you?) before reporting on whether it was distressing and if so, the extent that it bothered them. From these data, Total (i.e., the sum of endorsed items) was used as a measure of psychotic-like experiences.

### Additional variables

Child Substance Initiation: Neuroimaging study visits were excluded from analyses once participants reported substance use initiation. Alcohol, nicotine, and cannabis use initiation variables were derived based on endorsement of substance use at any yearly substance use interview or mid-year substance phone interview included in ABCD release 4.0. Specifically, alcohol initiation was defined as endorsing a full drink containing alcohol, outside of the context of religious ceremonies; nicotine initiation was defined as “more than a puff” in any form including tobacco cigarettes or cigars, e-cigarettes, hookah or pipes, or use of smokeless tobacco or chew, or nicotine patches, outside of the context of religious ceremonies; and cannabis initiation was defined as “more than a puff” in any form including smoking or vaping flower, oils, or concentrates, smoking blunts, or consuming edibles or tinctures, but not including synthetic cannabis or cannabis-infused alcoholic drinks.

Covariates were coded as in prior studies^19–21^. In brief:

#### Pubertal status

Parents and children both completed a 5-item scale on the child’s pubertal development^40^, combined to a summary score. The parent rating was used as the primary measure. The child rating was used if the parent rating was unavailable.

#### Child Race

Parents/caregivers selected from 26 categories. Dichotomous groups were formed for the most prevalent categories of race (i.e., White, Black, Asian, Pacific Islander, Native American) with remaining participants being assigned to Other. All variables were dummy coded as non-mutually exclusive dichotomous variables; as such, participants could be coded within more than one category.

#### Child Ethnicity

Parents/caregivers reported whether they consider the child to be hispanic/latinx. Maternal Education: Maternal education was recoded such that 12th grade, HS grad, and GED =12 years; some college and associate’s degree = 14 years; Bachelor’s degree = 16 years; Master’s degree = 18 years; Professional and Doctoral degrees = 20 years.

#### Household Income

Due to low endorsement of the first five of 10 household income levels, this variable was recoded such that the first five categories were assigned a value of one (i.e., <$50,000). The subsequent categories used were coded as two ($50,000-$74,999), three ($75,000-$99,999), four ($100,000-$199,999), and five ($200,000 or more), respectively.

#### Prenatal exposure to other substances

Child prenatal exposure to alcohol, tobacco, cocaine/crack, heroin/morphine, and oxycontin both before and after maternal knowledge of pregnancy were coded as separate dichotomous variables (cocaine/crack, heroin/morphine, and oxycontin were collapsed into a single variable due to low endorsement) based upon parent/caregiver retrospective report.

#### Additional covariates reported by parent/caregiver

Birth weight, family history of psychopathology (first-degree relative), unplanned pregnancy, maternal age at birth, use of prenatal vitamins, and gestational age when mother learned of pregnancy.

#### MRI covariates

scanner manufacturer and mean in-scanner motion during diffusion-weighted and resting state scans (not available for structural scans).

### Genetics

Genotyping, Quality Control, and Imputation: Saliva samples were genotyped on the Smokescreen array by the Rutgers University Cell and DNA Repository (now incorporated with other companies as Sampled; https://sampled.com/). The Rapid Imputation and COmputational PIpeLIne for Genome-Wide Association Studies (RICOPILI)^41^ was used to perform quality control (QC) on the 11,099 individuals with available ABCD Study phase 3.0 genotypic data, using RICOPILI’s default parameters (genotyping call rate >98%, inbreeding coefficient (F) < ±0.2, sex checks). The 10,585 individuals who passed these QC checks were aligned with broad self-identified racial groups using the ABCD Study parent survey. Of the 6,787 parents/caregivers indicating that their child’s race was only “white,” 5,561 of those individuals did not endorse any Hispanic ethnicity/origin. Next, variants were filtered to exclude those with missingness >2% and HWE p-value <1e-06. Using data from unrelated individuals (pi-hat ≤ 0.2) and an LD pruned set of common (MAF>0.05) and non-palindromic SNPs (and excluding MHC and chromosome eight inversion region), principal components analysis (PCA) was performed in RICOPILI using EIGENSTRAT^42^ to confirm the genetic ancestry of these individuals. This was done by merging the ABCD Study data with the 1000 Genomes reference panel, computing the mean and standard deviations for the 1000 Genomes ancestry populations for the top 6 PCs, and establishing that the previously identified group of 5,561 participants were genetically similar to the 1000 Genomes European panel by assessing whether they fell within 3 SDs of the mean for the top 6 PCs within this 1000 Genomes European reference population. After another round of QC on this subset, 5,556 European-ancestry individuals were retained. The European ancestry subset was then imputed to the TOPMed imputation reference panel16. Imputation dosages were converted to best-guess hard-called genotypes, and only SNPs with Rsq > 0.8, MAF > 0.01, missingness < 0.1, and HWE p-values >1E-06 were kept for PRS analyses.

#### Polygenic score

Polygenic scores (PGS) for cannabis use disorder (CUD) were generated using summary statistics from the latest CUD GWAS^43^. PGS were generated using PRS-CS^44^ (phi=default, iterations = 10,000, burn-in = 5,000), which uses bayesian regression and continuous shrinkage priors to create PGS that are competitive with state of the art methods.

### Statistical analyses

All outcome variables were winsorized (+/-3SDs) prior to analyses, and measures of psychopathology were log-transformed due to high skew. All variables were z-scored prior to analyses. Linear mixed-effect models were fit in R (v 4.1.5)^45^ using the ‘lme4’ package (v 1.1-34)^46^.

Linear mixed effect models testing the association of PCE with brain metrics included random intercepts for participant ID, site, and family ID, and fixed effects for Age, Age^2^, sex, pubertal status, familial relationship (i.e., twin, sibling), parental education and income, race, family history (i.e., first-degree relative) of drug, alcohol, or other mental health problems (n=6), prenatal exposure to alcohol, tobacco, or other drugs, scanner manufacturer, in-scanner motion (not available for structural scans), and the corresponding neuroimaging global variable (e.g., intracranial volume, mean fractional anisotropy of all white matter fibers, etc., except when testing said global variable as the outcome). Models were fit with and without the two PCE variables, and a log-likelihood ratio test was used to compare the two models. A significant p-value indicates that the addition of both PCE variables improves model fit, but does not give information on what drives the effect. We then examined the standardized regression effect size and associated confidence interval for the two PCE variables.

False discovery rate (FDR) correction for multiple comparisons was applied within each measure-type (e.g., structural volume only, white matter tract fractional anisotropy only, etc.). *Post-hoc* analyses confirmed that associations remained nominally significant with the inclusion of additional pregnancy-related covariates with higher missingness (i.e., mother’s age at the birth, birth weight, prenatal vitamins, planned pregnancy, when mother learned of pregnancy). Further *post-hoc* analyses 2) confirmed that the direction of association remained consistent when restricted to participants with genomically-confirmed European ancestry (N=4,962), which additionally included a polygenic score for cannabis use disorder and 10 principal components reflecting genomic ancestry as fixed effects.

Linear mixed effect models testing the association of brain metrics with psychopathology included random intercepts for participant ID, site, and family ID, and fixed effects for Age, Age^2^, sex, pubertal status, familial relationship, scanner manufacturer, in-scanner motion, and the corresponding neuroimaging global variable. False discovery rate (FDR) correction for multiple comparisons was again applied within each measure-type. Follow-up analyses included the full set of covariates used in primary PCE analyses (see above), to confirm that associations remained nominally significant with these covariates.

Mediation analyses were performed in R with the ‘mediation’ package (v 4.5.0)^35^. Analyses tested whether brain metrics at baseline mediated the association of PCE with psychopathology at the 1-year follow-up, and whether brain metrics at the 2-year follow-up mediated the association of PCE with psychopathology at the 3-year follow-up. As the ‘mediation’ package only accepts one random intercept, which needed to be a random intercept for Participant ID, study-site was modeled as 20 dummy-coded fixed effects, instead of a random effect. Further, only one individual from each family was included (chosen at random, unless there was at least one family member who was part of a PCE group), so that a random intercept for Family ID was not needed. As the package does not allow for categorical treatment variables with more than two levels in a mixed effect model, PCE was coded as a binary variable indicating any prenatal exposure. This was deemed reasonable as *post-hoc* tests in the significant brain metrics indicated that effects were directionally consistent between the two PCE groups (**Figure 2**). *Post-hoc* analyses were run for each PCE group separately to derive a separate estimate for each group. The mediation effect size reflects the expected effect if the association between PCE and psychopathology were fully mediated by the brain metric. FDR correction for multiple tests was applied across all analyses. *Post-hoc* analyses confirmed that associations remained nominally significant with: 1) the inclusion of additional pregnancy-related covariates with higher missingness (i.e., mother’s age at the birth, birth weight, prenatal vitamins, planned pregnancy, when mother learned of pregnancy) and 2) when analyses were restricted only to the baseline wave of collection. Further *post-hoc* analyses 3) confirmed that the direction of association remained consistent when restricted to participants with genomically-confirmed European ancestry (N=4,962), which additionally included a polygenic score for cannabis use disorder and 10 principal components reflecting genomic ancestry as fixed effects.

