## Supplement for "Prenatal cannabis exposure is associated with localized brain differences that partially mediate associations with increased adolescent psychopathology"

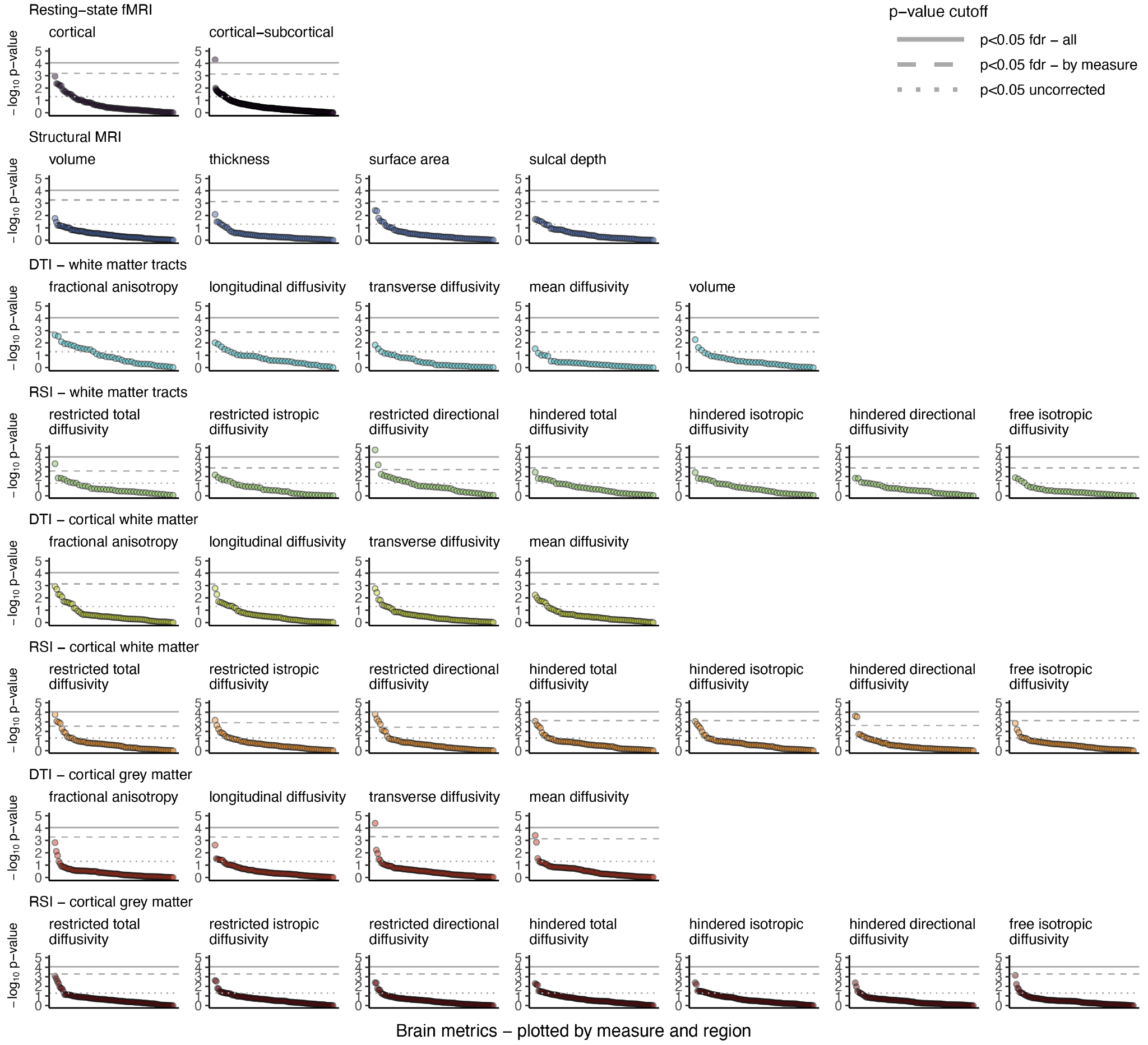

**Figure 1.** Association of prenatal cannabis exposure with brain metrics. Plots show the -log10 (negative base-10 logarithm) of the p-value for the association between brain metrics and prenatal cannabis exposure (PCE) from mixed effect regressions. Each imaging modality is plotted in a separate plot, with regions ordered within each plot from most to least significant. Horizontal lines are placed at p=0.05 uncorrected (dotted gray line), p<0.05 fdr-corrected for all comparisons of a given measure (i.e., all metrics within one plot; dashed gray line), and p<0.05 fdr-corrected for all 2,907 tests (solid gray line). **Figure 2** lists all regions surviving fdr-correction for comparisons of a given measure. Additional statistical information is provided in the **Supplemental Data File**.

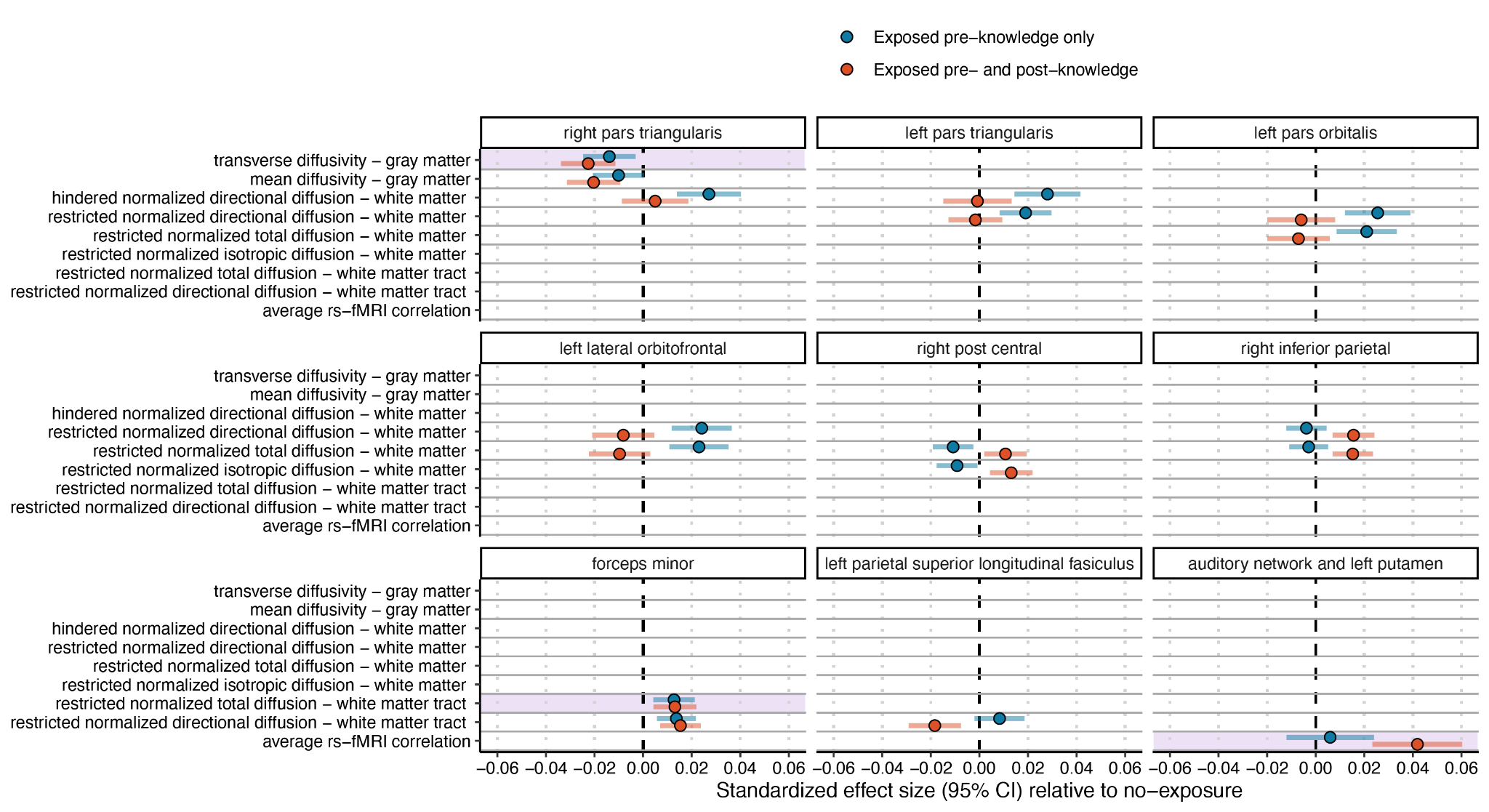

**Figure 2.** Significant associations of prenatal cannabis exposure with brain metrics, by exposure group. Standardized regression β effect sizes and 95% CIs from mixed-effect regressions assessing the association of prenatal cannabis exposure prior to maternal knowledge of pregnancy or prior to and post-maternal knowledge of pregnancy compared with no exposure. Nonsignificant outcomes are not shown. Y-axis reflects the measure-type, and each plot shows a single brain region to highlight overlap across different measures. Note that not all regions are measured with all modalities. Additional statistical information is provided in the **Supplemental Data File**. Regions surviving FDR multiple-test correction for all 2,907 tests are highlighted in purple.

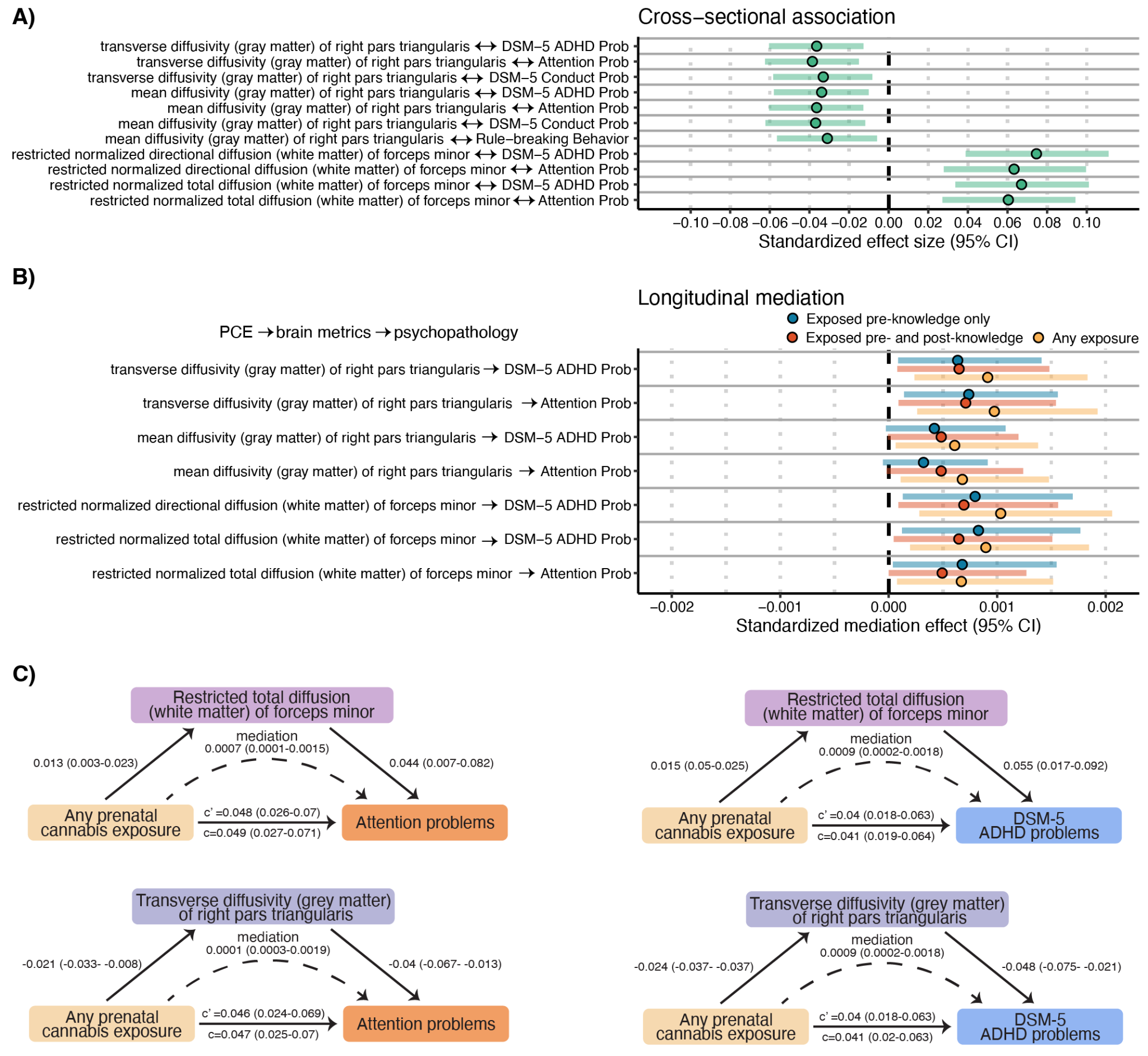

**Figure 3.** Association of brain metrics with childhood psychopathology. **A**) Standardized regression β effect sizes and 95% CIs from mixed-effect regressions assessing the association of significant (pfdr<0.05) brain metrics with psychopathology. **B**) Standardized mediation effect and 95% quasi-Bayesian CIs from longitudinal mediation analyses testing whether significant brain metrics mediate the association of prenatal cannabis exposure (PCE) with psychopathology. Analyses were run first collapsing across PCE groups (i.e., any exposure) and *post-hoc* analyses computed effects for each group separately. **C**) Mediation path diagrams for significant mediations (p<0.05 fdr) with regions which survived correction for all comparisons in primary analyses (**Figure 2**; restricted total diffusion of the forceps minor and transverse diffusivity of the right pars triangularis grey matter). Values reflect standardized effects and 95% CIs. c = total effect, c’ = direct effect.
